# Relationship between understanding of gender and LGBTQ and brain health with a focus on the triple network

**DOI:** 10.1101/2023.06.05.23290953

**Authors:** Taiko Otsuka, Keisuke Kokubun, Maya Okamoto, Yoshinori Yamakawa

## Abstract

Various studies have been conducted mainly in the fields of social sciences to address the lack of understanding of diversity in Japan regarding gender, LGBTQ, etc., but little progress has been made in promoting diversity in society. In this study, we aimed to clarify the specific brain states of people who have a high understanding of diversity (gender and LGBTQ) using the gray matter brain healthcare quotient (GM-BHQ), a brain health index obtained by MRI image analysis, and the results of a psychological questionnaire on diversity. As a result of the analysis, the GM-BHQ of the Central Executive Network (CEN) tended to be significantly higher than the estimated values from age, gender, and BMI in the group with a high understanding of gender equality. GM-BHQ of the Salience Network (SN) also tended to be relatively high. In addition, the GM-BHQ of SN, default mode network (DMN), and CEN, as well as whole brain GM-BHQ were significantly higher among those with a high understanding of LGBTQ than the estimated values. These results suggest that understanding diversity requires a healthy brain centered on the triple network that governs rational judgment, emotional adjustment, recognition of others, self-recognition, and value judgment of behavior.

## Introduction

Japan ranks 115th out of 146 countries (2022) in the “Gender Gap Index” released annually by the World Economic Forum, the lowest level among developed countries. Also, among Asian countries, the results were lower than those of South Korea, China, and ASEAN countries. This lack of diversity in Japanese society is viewed as a problem in the international community. In addition, Japan is the only G7 country that does not give legal rights to same-sex couples as a country and does not have anti-discrimination provisions regarding LGBTQ. Therefore, attention is focused on discussions at the G7 summit to be held in Japan in 2023. In response to this trend, the Liberal Democratic Party approved the “LGBT Understanding Promotion Bill” in May 2023, but the wording of the original bill has been revised in a rather conservative way. This is seen as a symbolic event that shows that the nation is not yet mature enough to accept diversity.

There have been many discussions on how to accept diversity in the social sciences such as economics and sociology. On the other hand, in the field of brain science, research on the differences in brain structure and function between men and women and LGBTQ people has been actively conducted (eg García, 2021; Lanz and Brown, 2019). In contrast, little research has been done on what the brains of people who accept such diversity are like. However, compassion is necessary to realize a society in which women and LGBTQ people are active, and compassion is said to be the ability to empathize with other people’s suffering (Rodríguez-Nieto et al., 2022). Therefore, it is not impossible to hypothesize and analyze the relationship between brain health and understanding of diversity by referring to the mechanisms of empathy that have been dealt with in the field of brain science. Here, essential to dealing with empathy is the triple network model (Menon, 2019), which consists of three networks: the Salience Network (SN), the Central Executive Network (CEN), and the Default Mode Network. (DMN).

Of these, the CEN, composed of the dorsolateral prefrontal cortex and the posterior parietal cortex, is important for the active retention and manipulation of information in working memory, attention, problem-solving, decision-making, and self-awareness (Fox et al., 2007; Goldberg et al., 2006; Koechlin and Summerfield, 2007; Miller and Cohen, 2001; Owen et al., 2005; Smith and Jonides, 1998; Wager and Smith, 2003). The SN is also a network that includes the ventrolateral prefrontal cortex (VLPFC) and the anterior insula (collectively referred to as the frontal insular cortex FIC) and the anterior cingulate cortex (ACC) (Seeley et al., 2010), responding to subjective degrees of salience, whether cognitive, homeostatic, or emotional (Goulden et al., 2014). The SN also acts as a switch between the CEN and the DMN, inhibiting the latter and activating the former when a salient stimulus or cognitive task is at hand, a process essential for attention and flexible cognitive control (Cai et al., 2015; Chen et al., 2015; Ham et al., 2013; Menon and Uddin, 2010; Sridharan et al., 2008; Uddin, 2015).

On the other hand, the DMN includes the medial posterior cortex, including the posterior cingulate cortex (PCC) and part of the frontal bone, the medial prefrontal cortex (MPFC), and the posterior temporal region around the temporoparietal junction (TPJ), including the inferior parietal lobule (IPL) (Andrews-Hanna et al., 2010; Buckner et al., 2008; Shulman et al., 1997). The DMN is preferentially activated when the individual is not focused on the external environment (Buckner et al., 2008) and is involved in various areas of cognitive and social processing. That is, the medial prefrontal cortex (MPFC) plays an important role in the social understanding of others, and the connections between the anterior MPFC (aMPFC) and the posterior and anterior cingulate cortices primarily contribute to self-other discrimination. On the other hand, the relationship between the dorsal MPFC (dMPFC) and the temporoparietal junction (TPJ) is mainly related to understanding the mental state of others (Li et al., 2014).

Interactions between these networks are related to everyday self-regulation (Krönke et al., 2020) and empathy for others (Acevedo et al., 2014; Bilevicius et al., 2018; Kim et al., 2017; Nummenmaa et al., 2008; Shamay-Tsoory et al., 2009). However, to the best of our knowledge, no research deals with the relationship between brain structure and function centered on these triple networks and understanding of diversity such as gender and LGBTQ. Therefore, in this study, by combining the GM-BHQ, which is a brain health index, and the psychological index related to acceptance of diversity, we investigated the structure of the brain centered on the triple network of people who accept diversity and those who do not. We would like to clarify the above differences and contribute to the development of diversity research in brain science and social science.

## Method

In this study, to clarify the differences in the acceptability of diversity and brain structure, we obtained a psychological index related to diversity and a brain health index, GM-BHQ, obtained by MRI image analysis. GM-BHQ was acquired between October and November 2023 after the approval by the Tokyo Institute of Technology’s ethical committee for “Brain information cloud (research ethics review committee for human subjects: permission number 2019007)”.

### Participants

We contacted 113 participants from whom we had acquired MRI data before by e-mail from April to May 2023. After that, we voluntarily asked them to cooperate with answering a questionnaire about diversity. There were 22 people (M: 20 people, F: 2 people) who participated in the questionnaire, and the average age was 47.6±12.7 years old.

### Questionnaire items

Gender and LGBTQ questions from the World Value Survey WAVE7 (Haerpfer, C. et al., 2022), which collects information on the values and beliefs of people around the world were extracted. As gender often raises issues in political and economic contexts (Elias and Rai, 2019), two related questions were used. Regarding LGBTQ, we considered that one of the three questions, “Same-sex couples can be good parents in the same way as heterosexual couples,” was related to the legal system of the country and deviated from the purpose of this study, which focused on individual acceptance. Therefore, we used two question items other than this question. Below are the four questions that were finally selected.

#### Gender

i. To what extent do you agree or disagree with the following opinions? “In general, men are better suited as political leaders than women.” 1: Strongly agree to 4: Strongly disagree, 5: Don’t know (Q29)
ii. How much do you agree or disagree with the following opinions? “When a wife earns more than her husband, problems always arise.” 1: Strongly agree - 5: Strongly disagree, 6: Don’t know (Q35)

#### LGBTQ

(iii) Are there any of the following people you wouldn’t want in your neighborhood? [Homosexual] 1: I don’t want to live in the neighborhood ∼ 6: I can live in the neighborhood (Q22)
(iv) What do you think about homosexuality? Do you think it is completely correct (acceptable) or completely wrong (not acceptable)? 1: Completely wrong (not acceptable) to 10: Completely correct (acceptable) (Q182)

Regarding questions (iii) and (iv), we have revised the question-and-answer items of the World Values Survey in line with this research. Specifically, for question (iii), the World Values Survey provides two responses: “1: I do not want to live in the neighborhood” and “2: It is OK to live in the neighborhood.” To extract more detailed differences in acceptability, this study was set with a 6-point method. In addition, regarding question (iv), since the World Values Survey has multiple response items including homosexuality, the question was “What do you think about each of the following?” However, in this study, as only homosexuality was extracted, the question content was adjusted as above.

### Calculation of Δ*GM-BHQ*

GM-BHQ is a standardized index of brain gray matter volume calculated from T1-weighted images with an average of 100 and a standard deviation of 15 (Nemoto et al., 2017). It has been approved as an international standard by the standardization organization ITU-T as a “numerical index representing the physical characteristics of the brain that indicate health-related conditions” (approval number: ITU-TH.861.0). See Nemoto et al. (2017) for details of the GM-BHQ estimation method.

Previous studies have shown that GM-BHQ can be predicted by multiple regression using age, gender, and BMI as variables (Nemoto et al., 2017). Based on this, we created a multiple regression equation with age, gender, and BMI as independent variables and GM-BHQ as dependent variables from the data of 113 people, and calculated the difference between the predicted GM-BHQ value from the equation and the actual GM-BHQ value, that is, ΔGM-BHQ. By using ΔGM-BHQ, we compare participants’ brain health and gender-LGBTQ understanding controlling for the effects of age, gender, and BMI.

Previous studies have shown that GM-BHQ at the whole-brain level was positively correlated with curiosity and perseverance (Kokubun et al., 2020), behavioral activation and empathic concern, and self-monitoring (Kokubun et al., 2022), and cognitive ability (Watanabe et al., 2021), and negatively correlated with fatigue and stress (Kokubun et al., 2018), an unbalanced diet (Kokubun and Yamakawa, 2019), and an unhealthy lifestyle (Kokubun et al., 2021). Longitudinal studies have shown that olfactory training (Watanabe et al., 2023) enhances GM-BHQ.

### Analysis method

This study tested the hypothesis that people who are more inclusive about gender and LGBTQ have higher GM-BHQ values. To that end, we first selected people who showed a high degree of acceptance regarding gender and LGBTQ in the four-questionnaire shown above. Specifically, regarding questions on gender, we extracted the respondents who chose either “4: Strongly Disagree” or “3: Disagree” for question i, and “5: Strongly Disagree” or “4: Disagree” for question ii. Regarding LGBTQ, we extracted those who selected “6: It’s OK to live in the neighborhood” for question iii and “10: Completely correct (acceptable)” for question iv. Next, a one-sample t-test was performed to confirm whether the mean value of ΔGM-BHQ in the extracted group with high acceptability was greater than zero. As evaluation targets, in addition to whole brain ΔGM-BHQ, we analyzed regional ΔGM-BHQ (CEN, SN, DMN), which are subscales of GM-BHQ.

## Result

Of the 22 participants in this study, one had type 1 diabetes. Because several previous studies have shown an association between diabetes and brain atrophy (Hansen et al., 2022; Ferguson, S. C., et al., 2005), the data for this one subject were excluded. Based on the method above, people with a high degree of gender and LGBTQ receptivity were extracted, 17 people were in the highly receptive group for gender and 9 people were in the highly receptive group for LGBTQ. In the figure 1, the results of a one-sample t-test were shown on the average ΔGM-BHQ values for the whole brain and for each region in these high receptive groups.

**Figure 1.**
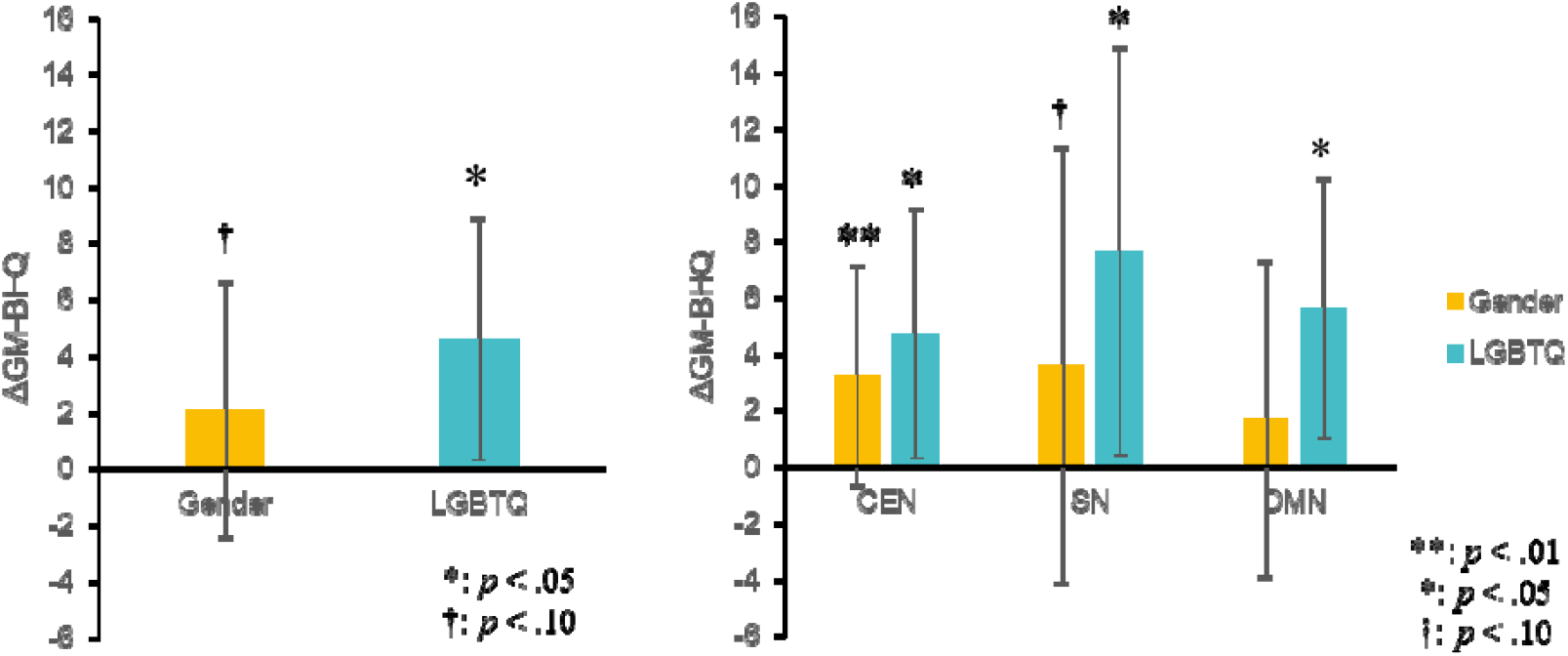
GM-BHQ and Gender/LGBTQ Awareness (Whole brain on the left, subscale on the right)

### Whole Brain BHQ (overall brain health)

For ΔGMBHQ in the whole brain, the gender-high receptivity group had an average value of 2.09 (SD = 4.50), which was not significant but relatively higher than 0 (t = -1.86, p = 0.08, Cohen’s d = 0.46). In addition, in the LGBTQ highly receptive group, the mean value (SD = 4.29) was significantly higher than the baseline value of 0 (t = -3.03, p = 0.02, Cohen’s d = 1.07).

### Region-specific BHQ (brain health level by region)

ΔGM-BHQ by site showed a level significantly higher than 0 only in the CEN in the gender receptive group, with an average value of 3.25 (SD = 3.90) (t= -3.33, p= 0.004, Cohen’s d = 0.83). Mean values were 3.62 (SD = 7.72) and 1.71 (SD = 5.61) in SN and DMN, respectively (SN: t = -1.87, p = 0.08, Cohen’s d = 0.47, DMN: t = -1.22, p = 0.242, Cohen’s d = 0.30). On the other hand, in the LGBTQ highly receptive group, ΔGMBHQ was significantly higher than 0 in all three regions of CEN, SN, and DMN, with mean values of 4.73 (SD = 4.45), 7.63 (SD = 7.24), and 5.60 (SD = 7.24), respectively. 4.63) (CEN: t = 3.00, p = 0.020, Cohen’s d = 1.06, SN: t = -2.98, p = 0.020, Cohen’s d = 1.05, DMN: t = -3.42, p = 0.011, Cohen’s d = 1.21).

## Discussion

This study showed that people with a high understanding of gender equality had a significantly higher CEN and a relatively higher SN GM-BHQ than the values estimated from age, gender, and BMI. On th other hand, it was shown that not only CEN, but also SN, DMN, and whole-brain GM-BHQ were significantly higher than the estimated values for those with a high understanding of LGBT. This can be said to indicate that the health level of the CEN area related to cognitive control is important to accept both gender and LGBT. The CEN region is a brain region related to working memory and rational judgment, such as the dorsolateral prefrontal cortex and the superior parietal lobule, and it is thought that these brain functions need to work soundly for a good understanding of gender (Fox et al., 2007; Goldberg et al., 2006; Koechlin and Summerfield, 2007; Miller and Cohen, 2001; Owen et al., 2005; Smith and Jonides, 1998; Wager and Smith, 2003).

In addition, the analysis results show that, especially for LGBTQ people, in addition to the CEN, the health level of the SN, which is responsible for the external monitoring function, and the DMN, which is closely related to sociality, is important. The SN region, which includes the insular cortex, anterior cingulate gyrus, and paracingulate gyrus, is a brain region associated with emotional response and regulation (Cai et al., 2015; Chen et al., 2015; Goulden et al., 2014; Ham et al., 2013; Menon and Uddin, 2010; Sridharan et al., 2008; Uddin, 2015). Therefore, it can be said that brain health centered on the SN, which appropriately regulates emotions, is required to reduce prejudice against LGBTQ people. On the other hand, the DMN region includes the posterior cingulate gyrus, medial superior frontal gyrus, precuneus, angular gyrus, and orbital inferior frontal lobe (Andrews-Hanna et al., 2010; Buckner et al., 2008; Li et al., 2014; Shulman et al., 1997). This suggests that more than gender equality, fair attitudes toward LGBTQ people require more complex social judgments and, therefore, brain health centered on the DMN, which is related to sociality.

A previous study performed an independent component analysis (ICA) on resting brain activity and showed that emotional empathy scores correlated with functional connectivity in the CEN, SN, and DMN (Bilevicius et al.., 2018). This result is consistent with empathy responses in previous studies using fMRI (Acevedo et al., 2014; Kim et al., 2017; Nummenmaa et al., 2008; Shamay-Tsoory et al., 2009), showing that emotional empathy involves movement, attention, and self-referential processing. Here, affective empathy refers to the ability to understand, infer, judge, and share the emotional experiences of others (Gallese, 2003; Shamay-Tsoory et al., 2009), and is important in social interaction (Baron-Cohen et al., 2001, Baron-Cohen and Wheelwright, 2004). As such, previous studies have shown that deficits in emotional empathy are associated with alcoholism (Maurage et al., 2011) and social anxiety disorder (Morrison et al., 2016).

Previous research also indicates that CEN, SN, and DMN are uniquely associated with empathy. fMRI experiments showed that both empathetic and permissive judgments activate the superior frontal gyrus that spans the CEN and DMN (Farrow et al., 2001). The superior frontal gyrus is thought to contribute to cognitive functions such as self-awareness (Goldberg et al., 2006) and working memory (WM) in conjunction with the action of the sensory system (Alagapan et al. 2019; Boisgueheneuc et al., 2006). This suggests that empathy activates specific brain regions and contributes to social cohesion (Farrow et al., 2001). Similarly, studies on racial bias show that activation of regions of the frontal cortex associated with control and regulation modulates amygdala activity, thereby suppressing racist emotions. (Banaji et al., 2008). Also, some fMRI studies suggest that the lack of empathy is primarily due to a defective SN switching function. Dysfunctional SNs are involved in the activation of the DMN, which leads to self-focused attention and empathy disorders such as narcissism (Jankowiak-Siuda and Zajkowski, 2013). Furthermore, the AIs and dACCs that make up the SN have been associated with empathy themselves (Fallon et al., 2020; Gu et al., 2010; Timmers et al., 2018).

Alternatively, data-driven quantitative reasoning studies suggest that SN and CEN facilitate individuals to build long-term social relationships through emotional processing and cognitive control and that DMN facilitates individuals to develop long-term social relationships through mentalizing processes. It has been suggested that it predicts the experiences, beliefs, and intentions of others and facilitates interaction (Li et al., 2022). In particular, DMN has been implicated in both functional and structural studies as prosocial personality traits such as extraversion and agreeableness (Mars et al., 2012; Sampaio et al., 2014; Coutinho et al., 2013), and it is associated with self-cognitive empathy (Silva et al., 2018; Esménio et al., 2019). On the other hand, a recent systematic review shows that people with low compassion tend to have either low reward-related neuronal area activity or gray matter volume (Novak et al., 2022). Consistent with this, our previous studies have shown that whole-brain gray matter is positively correlated with psychological variables representing behavioral activation, empathic concern, and self-monitoring (Kokubun et al.., 2022).

This series of previous studies provides compelling reasons why CEN is required for understanding gender, and why a triple network consisting of CEN, SN, and DMN is required for understanding LGBTQ. First, women make up half of Japan’s population and are familiar to men. In a survey in which managers of companies with diversity management strategies were asked, “What are the benefits obtained by accepting diversity?”, the following effects were confirmed. “Acquisition of human resources”, “Improvement of performance”, “Enhancement of brand power and reputation”, “Vitalization of innovation”, etc. (Ministry of Economy, Trade and Industry, 2020). Alternatively, research on the relationship between women’s participation in parliament and the economy shows that a 10-percentage point increase in women’s representation in parliament leads to a 0.74 percentage point increase in GDP growth (Mirziyoyeva and Salahodjaev, 2023). Furthermore, items such as “degree of freedom of choice in life” and “tolerance” are pushing down the level of happiness in Japan. A study also found a statistically significant correlation (correlation coefficient 0.486 in 124 countries) between the gender gap index and subjective well-being. Therefore, it is an important time for business and political leaders to embrace diverse perspectives, not only for the social contribution reasons that have been thought of so far, but also to strengthen the profit structure of the economy at the corporate and national level, and to increase the happiness of the people. In this situation, increasing diversity is one of the rational decisions necessary to demonstrate high performance and increase happiness. Our results are convincing, showing that gender understanding was most strongly associated with CEN, which governs attention and decision-making.

On the other hand, it is said that 8.9% of the population of Japan is LGBTQ, and it is difficult to recognize them as a familiar existence. As a result, in the above survey results, the most common group in Japan is the “knowledgeable other people (34.1%): people who maintain the status quo who have knowledge but do not have the opportunity to feel a sense of challenge because they are not familiar with the person concerned.” Therefore, unfortunately, it is true that prejudice against LGBTQ still exists. This means that it is necessary to have the ability to “imagine and consider the feelings of minorities” to understand LGBTQ. In other words, it is thought that people with a high degree of understanding of LGBTQ have an inclusive mindset by simultaneously using their rational judgment and the ability to consider others who are not close to them. By the way, sympathy for LGBTQ by activating the triple network of CEN plus SN and DMN is extremely important in the realization of design thinking and art thinking, which have been important in the business world for about 10 years. Design thinking requires both the ability to design based on empathy for a variety of things and the ability to think logically to construct a business model. This is exactly the domain of the triple network. Therefore, judging from the results of this analysis, it can be surmised that people with a high degree of understanding of diversity are likely to have high potential as business people and managers.

This study is the first to show that brain health centered on the triple network is necessary for understanding diversity such as gender and LGBTQ, contributing to the development of diversity research in brain science and social science.

### Limitation

The small sample size casts doubt on the robustness of the results. Future studies should validate this study by performing similar studies with larger sample sizes. In addition, the cross-sectional analysis showed a correlation, not a causal relationship between variables. In the future, the method of longitudinal analysis should be adopted to verify the results of this study.

## Conclusion

In this study, analysis was performed using the brain health index GM-BHQ obtained by MRI image analysis and the results of a psychological questionnaire. As a result, people with a high understanding of diversity such as gender and LGBTQ had a healthy brain centered on the triple network consisting of CEN, SN, and DMN compared to the values estimated from age, gender, and BMI.

## Data Availability

All data produced in the present study are available upon reasonable request to the authors

